## Supplementary information for "Unraveling the drivers of leptospirosis risk in Thailand using machine learning"

### XGBoost hyperparameters

In our work, we tuned several key hyperparameters to optimize the XGBoost model's performance and prevent overfitting. The *n_estimators* parameter determines the number of trees grown for the classification task, while the *learning_rate* controls the rate at which the model learns and corrects errors for each new tree. The *max_depth* parameter sets the maximum depth of each tree, with deeper trees more likely to overfit the data and shallower trees (stumps) potentially underfitting. *Subsample* and *colsample_bytree* introduce randomness by subsampling the training data and feature columns, respectively, for each tree, reducing the likelihood of overfitting. The *min_child_weight* parameter sets the minimum number of samples required in each leaf node, further preventing overfitting. *Gamma* serves as a regularization parameter in the loss function, while *alpha* and *lambda* control the L1 and L2 regularization terms on the weights, respectively. Careful tuning of these hyperparameters is crucial to ensure optimal model performance and generalization to unseen data [1].

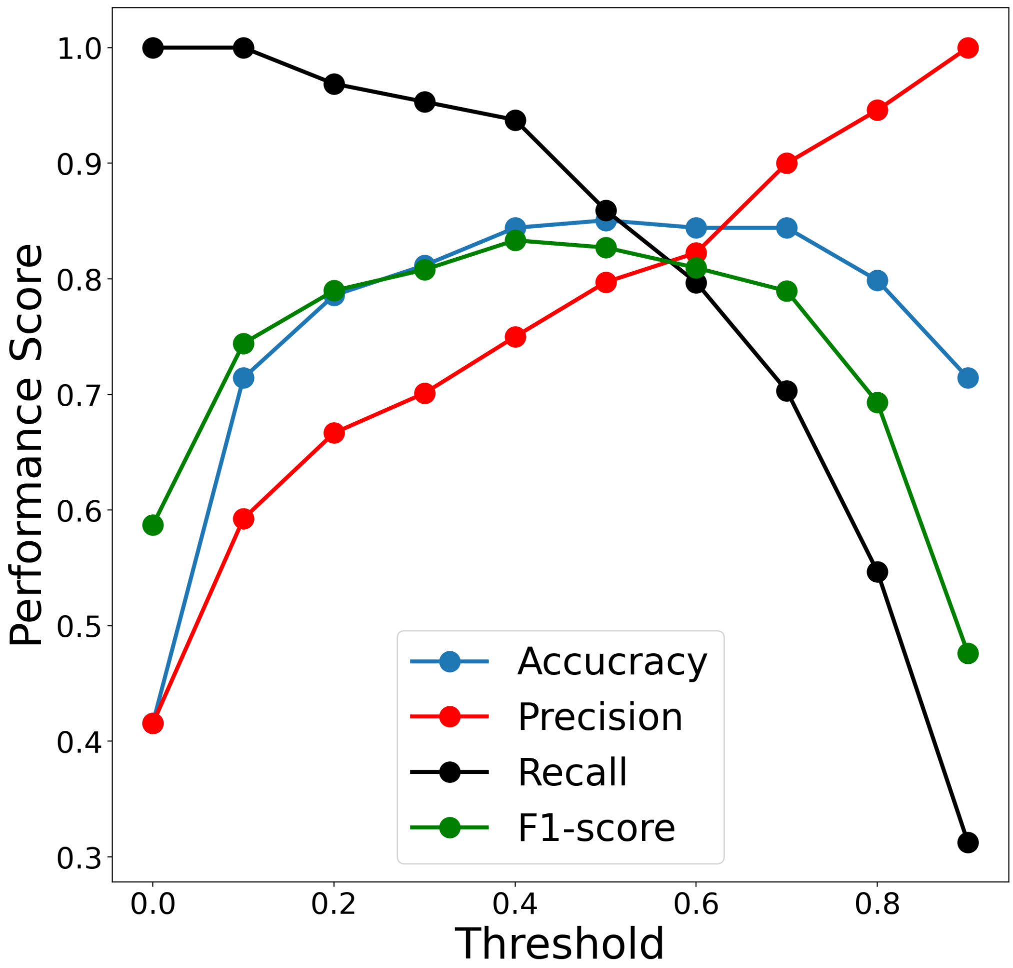


**Figure S1.** Threshold sensitivity analysis of model performance on test data (2018-2019). Performance metrics (accuracy, precision, sensitivity, and F1-score) are evaluated across different classification thresholds, where the threshold determines the cutoff between predicted low-risk (0) and high-risk (1) provinces. For each threshold value, provinces are classified as high-risk when their predicted probability exceeds the threshold.

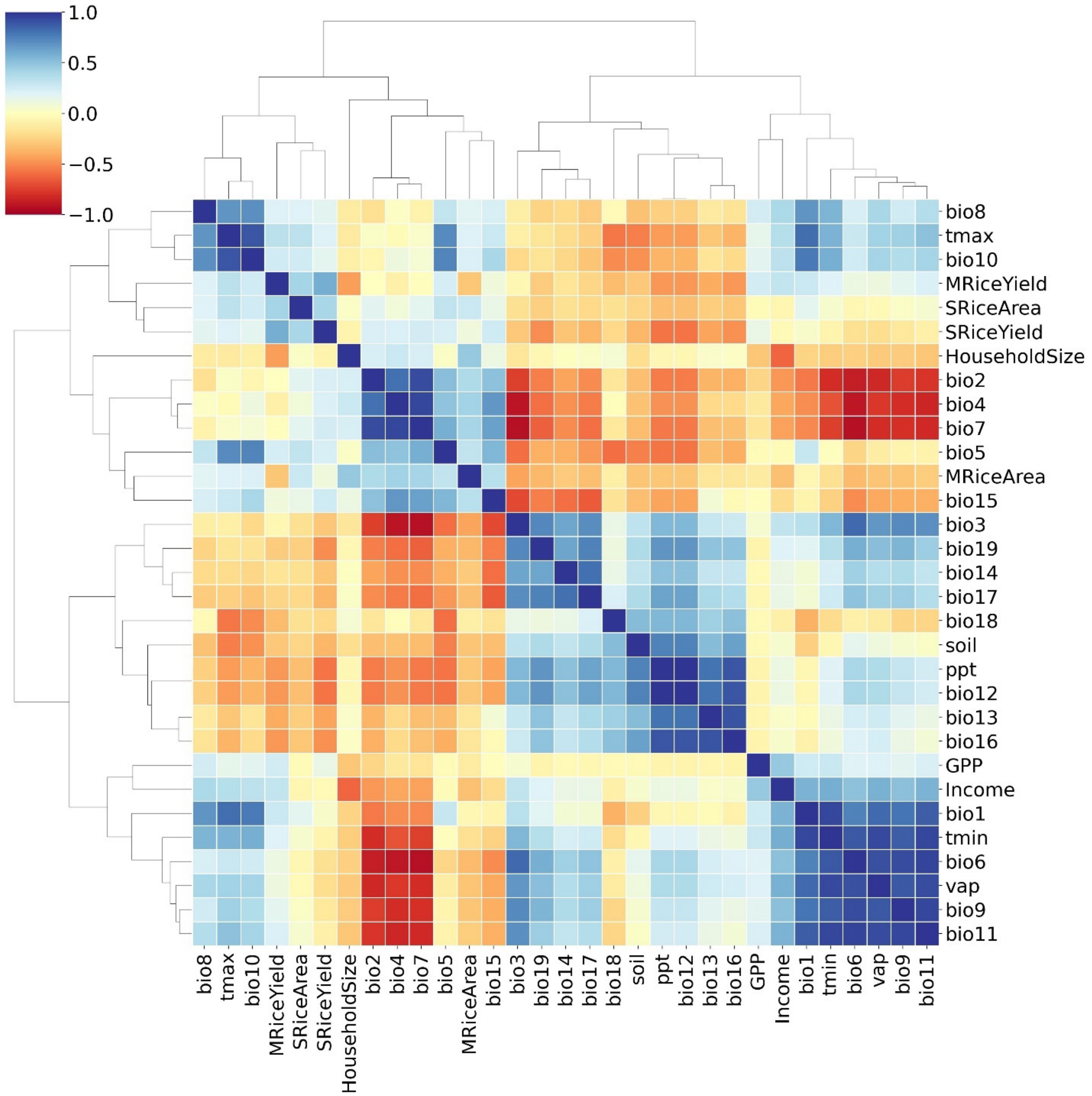


**Figure S2.** Hierarchically clustered correlation matrix of predictive features. The heatmap displays Pearson correlation coefficients between all model features, visualized using seaborn's clustermap function. Correlation strength and direction are indicated by color intensity (red = positive, blue = negative). Features are hierarchically clustered to reveal groups of highly correlated variables, highlighting potential multicollinearity in the dataset.

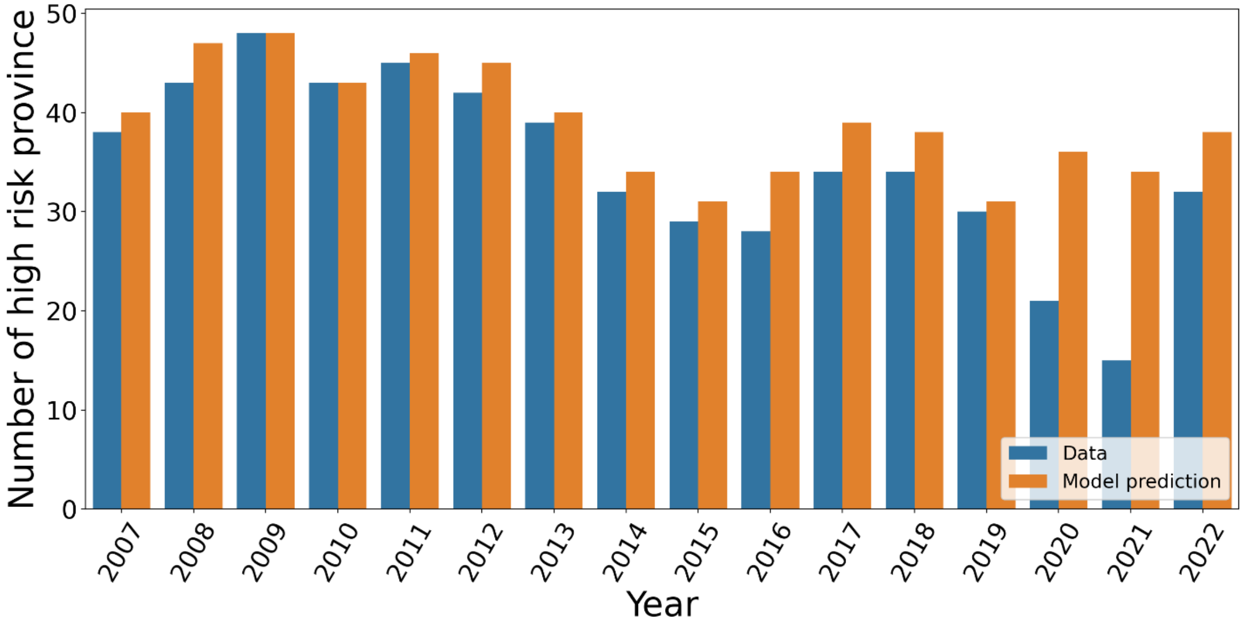


**Figure S3.** Temporal distribution of predicted high-risk provinces across pandemic periods. Comparison of model-predicted versus observed counts of high-risk provinces during pre-pandemic (2007-2019), pandemic (2020-2021), and post-pandemic (2022) periods. Using the model trained on 2007-2017 data, predictions reveal substantial changes in the number of high-risk provinces, particularly during the pandemic period. This visualization highlights the pandemic's potential impact on leptospirosis risk patterns and/or surveillance capabilities across Thailand's provinces.

### Reference

1. Chen T, Guestrin C. XGBoost: A Scalable Tree Boosting System. Proceedings of the 22nd ACM SIGKDD International Conference on Knowledge Discovery and Data Mining; San Francisco, California, USA: Association for Computing Machinery; 2016. p. 785–94.
